# Social and behavioral risk factors are associated with age-related differences in sustained attention: A cross-sectional GradCPT analysis of the All of Us dataset

**DOI:** 10.64898/2026.10.01.26364102

**Authors:** Natan Coresh-Chiappori, Theresa Boyer, A. Richey Sharrett, Ingrid K. Tulloch

## Abstract

**Background:** Attentional control declines with normal aging, yet the role of modifiable lifestyle factors in accelerating this decline remains incompletely characterised in large, diverse populations. We examined associations between four modifiable risk factors (smoking intensity, alcohol binge drinking, social satisfaction and blood pressure) and sustained attention performance in 13,866 adults drawn from the All of Us Research Program.

**Methods:** Cognitive performance was assessed using the Gradual-Onset Continuous Performance Task (GradCPT). A composite cognitive efficiency score was derived via principal component analysis of four GradCPT metrics: d′ (perceptual sensitivity), response criterion, median reaction time and reaction time variability. Associations between cognitive efficiency and each risk factor were examined using multiple linear regression adjusted for age, sex and race/ethnicity. Effect sizes were expressed as age-equivalent years of cognitive aging (age-equivalent years = regression coefficient for the risk factor divided by the regression coefficient for age) to aid clinical interpretation.

**Results:** GradCPT performance declined progressively with age (p < 0.001). Social satisfaction emerged as the strongest lifestyle predictor: low social satisfaction was associated with approximately 5.8 additional age-equivalent years of cognitive aging relative to high social satisfaction. Smoking showed a dose-response pattern, with heavy smokers (> 20 cigarettes/day) exhibiting an approximately 5-year age-equivalent deficit versus never-smokers. Stage 2 hypertension showed a modest, non-significant negative association. Binge drinking showed limited significant associations, likely complicated by survivor and abstainer biases. Bootstrapped analyses of combined exposures showed that combinations of risk factors, particularly those involving low social satisfaction, produced larger cognitive deficits than expected under an additive model.

**Conclusions:** These findings support an interactive rather than a purely additive model of lifestyle-related cognitive vulnerability and highlight the GradCPT as a subclinical measure of attention-based cognitive risk in non-clinical populations.

## Introduction

Cognitive aging involves domain-specific changes that emerge well before clinical thresholds are reached. Sustained attention, mediated by prefrontal and anterior cingulate circuits, is particularly sensitive to age-related neurodegeneration and is disrupted early in conditions such as Alzheimer’s disease [1]. The lifetime risk of dementia exceeds 42% after age 55 [2], yet current medicine offers only modest symptomatic relief [3, 4], making prevention through modifiable risk factor management a critical priority. Aging is further associated with structural prefrontal changes, namely dendritic spine loss, which reduces neural efficiency and plasticity; behavioral exposures may accelerate these changes [5].

The Gradual-Onset Continuous Performance Task (GradCPT) [6], selected as the cognitive measure in the All of Us Research Program, provides a sensitive behavioral measure of sustained attention and inhibitory control. Participants view a sequence of greyscale city and mountain images transitioning every 800 ms and are instructed to respond to city images (89.3% of trials) while withholding responses to mountain images (10.7%). Performance is quantified by d′ (signal detection sensitivity), response criterion (decision bias), median reaction time and reaction time variability. In a study of 10,430 participants, Fortenbaugh et al. [7] demonstrated that GradCPT performance peaks in the early 40s and declines thereafter, following a trajectory distinct from fluid or crystallised intelligence. These characteristics make the GradCPT well suited and age-sensitive for large-scale population studies.

Modifiable risk factors linked to cognitive decline include smoking, alcohol use, social isolation and hypertension. Longitudinal evidence indicates that smoking accelerates structural brain aging in proportion to exposure intensity [8], while social disengagement is associated with substantially faster cognitive decline; highly socially active individuals show nearly 70% slower decline than their least active counterparts [9]. Midlife hypertension accounts for 15–20% of dementia cases by age 80 [10]. These factors rarely act in isolation, and prior work suggests synergistic effects when risk behaviours co-occur [11, 12], although this has not been examined in the context of GradCPT performance.

The GradCPT was adopted by the All of Us Research Program as a language- and education-agnostic test of sustained visual attention with good psychometric properties [7]. However, no previous studies have examined GradCPT performance in relation to modifiable risk factors. The present study addresses these gaps using data from 13,866 participants in the All of Us Research Program to: (1) characterise GradCPT performance trajectories across the adult lifespan; (2) assess independent associations between four modifiable risk factors and age-adjusted cognitive efficiency; and (3) test whether combinations of risk factors produce synergistic cognitive deficits beyond what individual effects would predict.

## Methods

### Dataset and participants

Data were drawn from the All of Us Research Program (National Institutes of Health) [13], a large precision medicine initiative with over 849,000 enrolled participants as of March 2025. This study used the All of Us Registered Tier Dataset v8, available to authorized users on the Researcher Workbench. Of 24,864 individuals who completed the GradCPT “City or Mountain” task with valid performance data, 13,866 had complete survey responses for all four lifestyle risk factor domains and were included in the final analytic cohort (Figure 1). Quality control flags excluded records showing sustained non-response (≥ 75 consecutive withheld trials), a high omission error rate (> 50% of go-trials missed) or extended disengagement (> 60 s of flagged trials).

**Figure 1.**
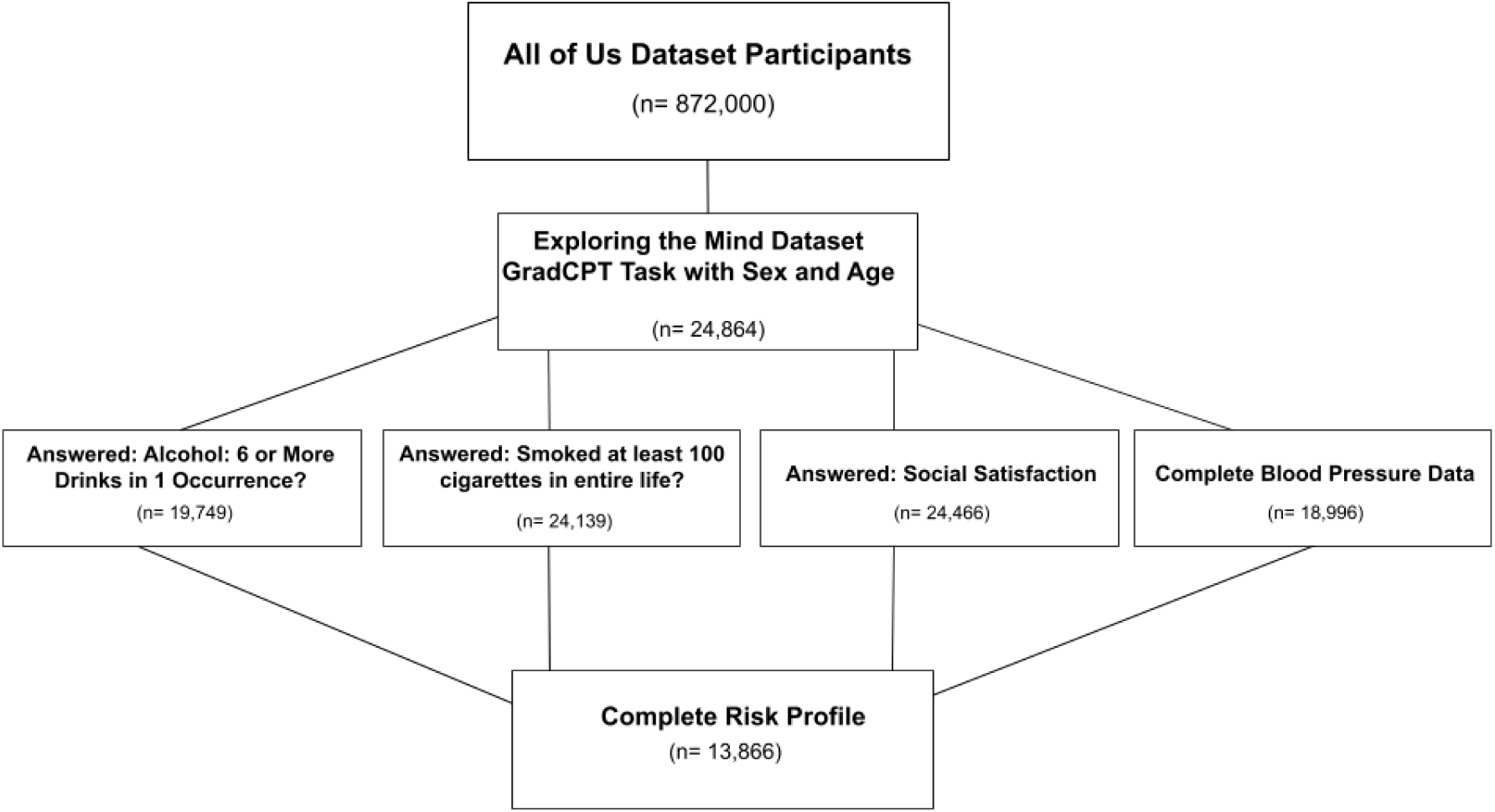
Participant selection flow diagram. Starting from all enrolled All of Us participants, the sample was restricted to those completing the GradCPT with valid performance data and available lifestyle, demographic and health variables. Final analytic cohort: N = 13,866.

### Measures

Smoking status was assessed by self-reported lifetime cigarette use and categorised as never smoker, ever smoker (≤ 10 cigarettes/day), ever smoker (11–20/day) or ever smoker (> 20/day). Alcohol binge drinking was assessed by past-year frequency of consuming ≥ 6 drinks on a single occasion and categorised as never (in the past year), less than monthly, or monthly or more. Social satisfaction was assessed using a single PROMIS item (“How would you rate your satisfaction with your social activities and relationships?”) and categorised as high, moderate or low. This item captures subjective satisfaction and should be interpreted as a distinct but relevant measure of sociability, a known risk factor in the cognitive aging literature. Blood pressure was classified from measured systolic (SBP) and diastolic (DBP) values as normal (SBP < 120 and DBP < 80 mmHg), elevated (SBP 120–129 and DBP < 80 mmHg), stage 1 hypertension (SBP 130–139 or DBP 80–89 mmHg) or stage 2 hypertension (SBP ≥ 140 or DBP ≥ 90 mmHg, or current antihypertensive medication use). The observation closest in time to the GradCPT assessment was used for all health measurements.

### Cognitive efficiency composite

Four GradCPT performance metrics (d′ [perceptual sensitivity], criterion [response bias], median reaction time and coefficient of variation of reaction time [attentional stability]) were combined into a single composite via principal component analysis (PCA). The first principal component (PC1) explained 57.6% of the total shared variance and was designated the cognitive efficiency score (Table 1). d′ contributed most strongly (loading = 0.55), reflecting accurate and timely target discrimination. Higher cognitive efficiency scores indicate better sustained attentional performance.

**Table 1.**
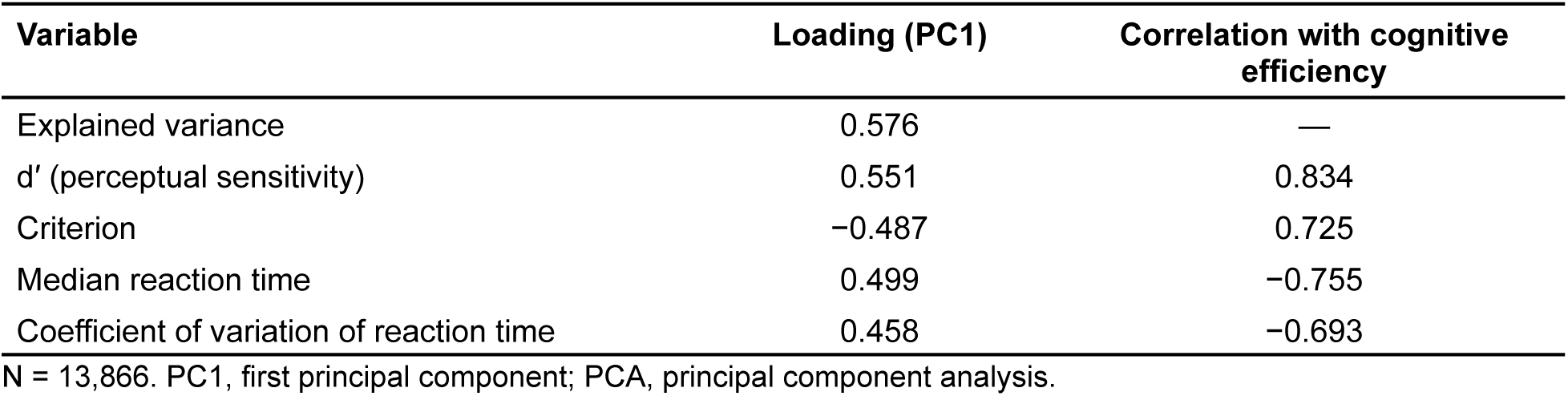
PCA loadings for the cognitive efficiency composite score.

| Variable | Loading (PC1) | Correlation with cognitive efficiency |
| --- | --- | --- |
| Explained variance | 0.576 | — |
| $d'$ (perceptual sensitivity) | 0.551 | 0.834 |
| Criterion | -0.487 | 0.725 |
| Median reaction time | 0.499 | -0.755 |
| Coefficient of variation of reaction time | 0.458 | -0.693 |

N = 13,866. PC1, first principal component; PCA, principal component analysis.

### Age-equivalent scaling

To express effect sizes in clinically interpretable terms, differences in cognitive efficiency were converted to age-equivalent years using the normative adult decline estimated from cubic regression splines fitted to the full sample. The predicted decline between ages 40 and 75 (−0.0179 cognitive efficiency units/year) served as the scaling denominator. A between-group difference of, for example, −0.068 units corresponds to −0.068 / −0.0179 ≈ 3.8 age-equivalent years of additional cognitive aging. Despite its interpretability, this measure has a limitation: the scaling factor was derived from cross-sectional age differences rather than true within-person decline, which the present data cannot confirm.

### Statistical analyses

Multiple linear regression (ordinary least squares, OLS) was used to assess independent associations between cognitive efficiency and each risk factor, adjusting for chronological age, sex and race/ethnicity. Natural cubic spline regression (knots at age tertiles) was used to model age trajectories stratified by each risk factor group. For analyses of combined exposures, a factorial OLS regression model included all 16 binary permutations of four risk indicators (heavy smoking, monthly-or-more binge drinking, low social satisfaction and high blood pressure), with full factorial interaction terms and HC3 heteroskedasticity-consistent standard errors. Bootstrap resampling (5000 iterations) generated confidence intervals (CIs) for multi-factor contrasts, with predictions anchored at age 50 to isolate combined-exposure effects from age-related variation. Sex was included as a covariate in all models. Analyses were conducted in a Python Jupyter Notebook. In accordance with the All of Us Data and Statistics Dissemination Policy, no participant counts between 1 and 20 are reported.

## Results

### Sample characteristics

The final analytic cohort comprised 13,866 participants (mean age 61.1 years, standard deviation [SD] 15.1; 67.1% female; 82.2% non-Hispanic White). The majority were never-smokers (65.7%), never binge drinkers (73.1%) and reported high social satisfaction (62.8%). One-third had normal blood pressure (33.3%), and 20.3% were classified as having stage 2 hypertension (Table 2).

**Table 2.** Characteristics of the study population.

| Characteristic | Category | n (%) | Mean (SD) |
| --- | --- | --- | --- |
| Total population | — | 13,866 (100%) | — |
| Sex | Male (ref) | 4562 (32.9%) | — |
|  | Female | 9304 (67.1%) | — |
| Age, years | — | — | 61.1 (15.1) |
| Age group, years | 18–29 | 509 (3.7%) | — |
|  | 30–39 | 1170 (8.4%) | — |
|  | 40–49 | 1429 (10.3%) | — |
|  | 50–59 | 2109 (15.2%) | — |
|  | 60–69 | 3742 (27.0%) | — |
|  | 70–79 | 4022 (29.0%) | — |
|  | ≥ 80 | 885 (6.4%) | — |
| Smoking status (lifetime) | Never smoker (ref) | 9109 (65.7%) | — |
|  | Ever smoker (≤ 10/day) | 2323 (16.8%) | — |
|  | Ever smoker (11–20/day) | 1756 (12.7%) | — |
|  | Ever smoker (> 20/day) | 678 (4.9%) | — |
| Alcohol binge drinking (≥ 6 drinks) | Never (ref) | 10,138 (73.1%) | — |
|  | Less than monthly | 2835 (20.4%) | — |
|  | Monthly | 579 (4.2%) | — |
|  | Weekly | 238 (1.7%) | — |
|  | Daily | 78 (0.6%) | — |
| Social satisfaction | High (ref) | 8703 (62.8%) | — |
|  | Moderate | 3401 (24.5%) | — |
|  | Low | 1762 (12.7%) | — |
| Blood pressure category | Normal (ref) | 4613 (33.3%) | — |
|  | Elevated | 2303 (16.6%) | — |
|  | Stage 1 hypertension | 4138 (29.8%) | — |
|  | Stage 2 hypertension | 2812 (20.3%) | — |

N = 13,866. ref, reference category; SD, standard deviation.

### Cognitive efficiency across age

Cognitive efficiency declined across adulthood (Figure 2), with estimated declines of −0.875 units between ages 20 and 50 and −1.073 units between ages 50 and 80. The average annual decline between ages 40 and 75 was −0.0179 cognitive efficiency units/year, used as the age-equivalent reference scale.

**Figure 2.**
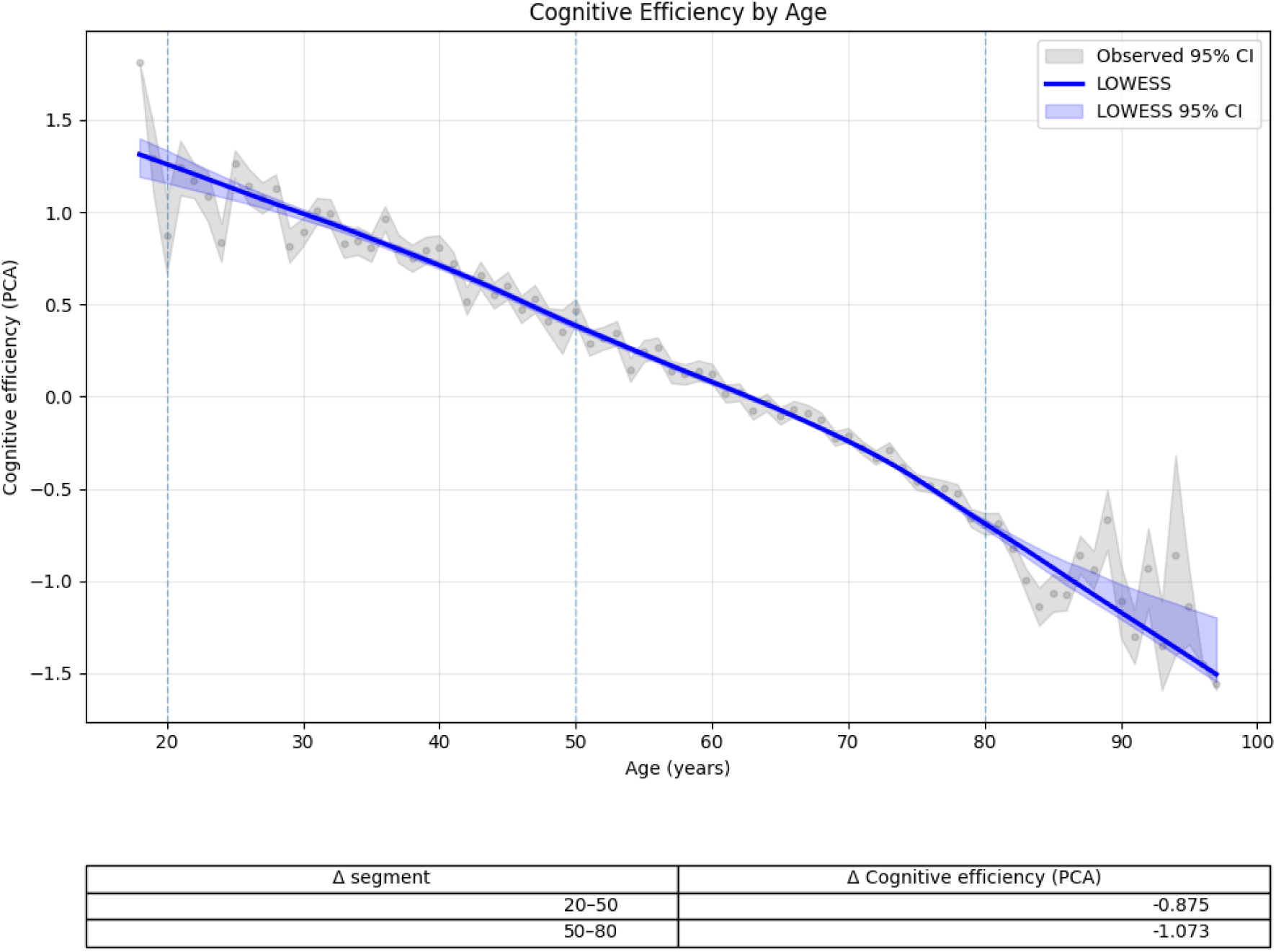
Cognitive efficiency across age. Cognitive efficiency (PC1) plotted across age using natural cubic regression splines with 95% confidence intervals. Vertical dashed lines denote spline breakpoints; the inset table summarises estimated declines across calibration age intervals.

### Regression results

Age was the dominant predictor of cognitive efficiency (β = −0.035 per year, p < 0.001; Table 3). Female sex was associated with lower cognitive efficiency relative to male sex (β = −0.249, p < 0.001; approximately 5.1 age-equivalent years). Non-Hispanic Black race/ethnicity showed the largest race-related difference (β = −0.411, p < 0.001; approximately 8.7 years). Among modifiable risk factors, social satisfaction showed the strongest effects: moderate satisfaction (β = −0.123, p < 0.001; +2.4 years) and low satisfaction (β = −0.191, p < 0.001; +3.6 years) relative to high satisfaction. Smoking showed a dose-dependent pattern: 11–20 cigarettes/day (β = −0.110, p < 0.001; +2.6 years) and > 20/day (β = −0.158, p < 0.001; +3.9 years), while ≤ 10/day was non-significant. Binge drinking showed a significant effect only for infrequent bingeing (less than monthly: β = −0.062, p = 0.001; +1.3 years); more frequent categories were not statistically significant. Blood pressure categories showed small, mostly non-significant associations, and stage 2 hypertension showed a modest negative association (β = −0.034, p = 0.108). The model explained approximately 26% of the variance in cognitive efficiency.

**Table 3.** Multiple linear regression of cognitive efficiency (PC1) on demographic and lifestyle risk factors.

| Risk factor | $\beta$ (SE) | t | p value |
| --- | --- | --- | --- |
| Age (years) | -0.035 (0.001) | -62.984 | < 0.001 |
| Sex: male | Ref | — | — |
| Sex: female | -0.249 (0.016) | -15.464 | < 0.001 |
| Smoking: never smoker | Ref | — | — |
| Smoking: ever smoker ($\leq 10$/day) | -0.027 (0.020) | -1.337 | 0.181 |
| Smoking: ever smoker (11–20/day) | -0.110 (0.023) | -4.774 | < 0.001 |
| Smoking: ever smoker ($> 20$/day) | -0.158 (0.035) | -4.532 | < 0.001 |
| Binge drinking: never | Ref | — | — |
| Binge drinking: less than monthly | -0.062 (0.019) | -3.228 | 0.001 |
| Binge drinking: monthly | 0.009 (0.037) | 0.228 | 0.820 |
| Binge drinking: weekly | -0.011 (0.057) | -0.200 | 0.841 |
| Binge drinking: daily | -0.120 (0.098) | -1.229 | 0.219 |
| Social satisfaction: high | Ref | — | — |
| Social satisfaction: moderate | -0.123 (0.017) | -7.051 | < 0.001 |
| Social satisfaction: low | -0.191 (0.023) | -8.375 | < 0.001 |
| Blood pressure: normal | Ref | — | — |
| Blood pressure: elevated | 0.017 (0.022) | 0.755 | 0.450 |
| Blood pressure: stage 1 hypertension | 0.008 (0.019) | 0.428 | 0.668 |
| Blood pressure: stage 2 hypertension | -0.034 (0.021) | -1.609 | 0.108 |

### Sex differences

Males showed higher baseline cognitive efficiency but a steeper decline with age relative to females (Figure 3). This is consistent with prior work documenting that males outperform females at baseline on visuospatial tasks but exhibit faster longitudinal decline [14]. The sex difference corresponded to approximately 7 age-equivalent years at midlife in the age-anchored model.

**Figure 3.**
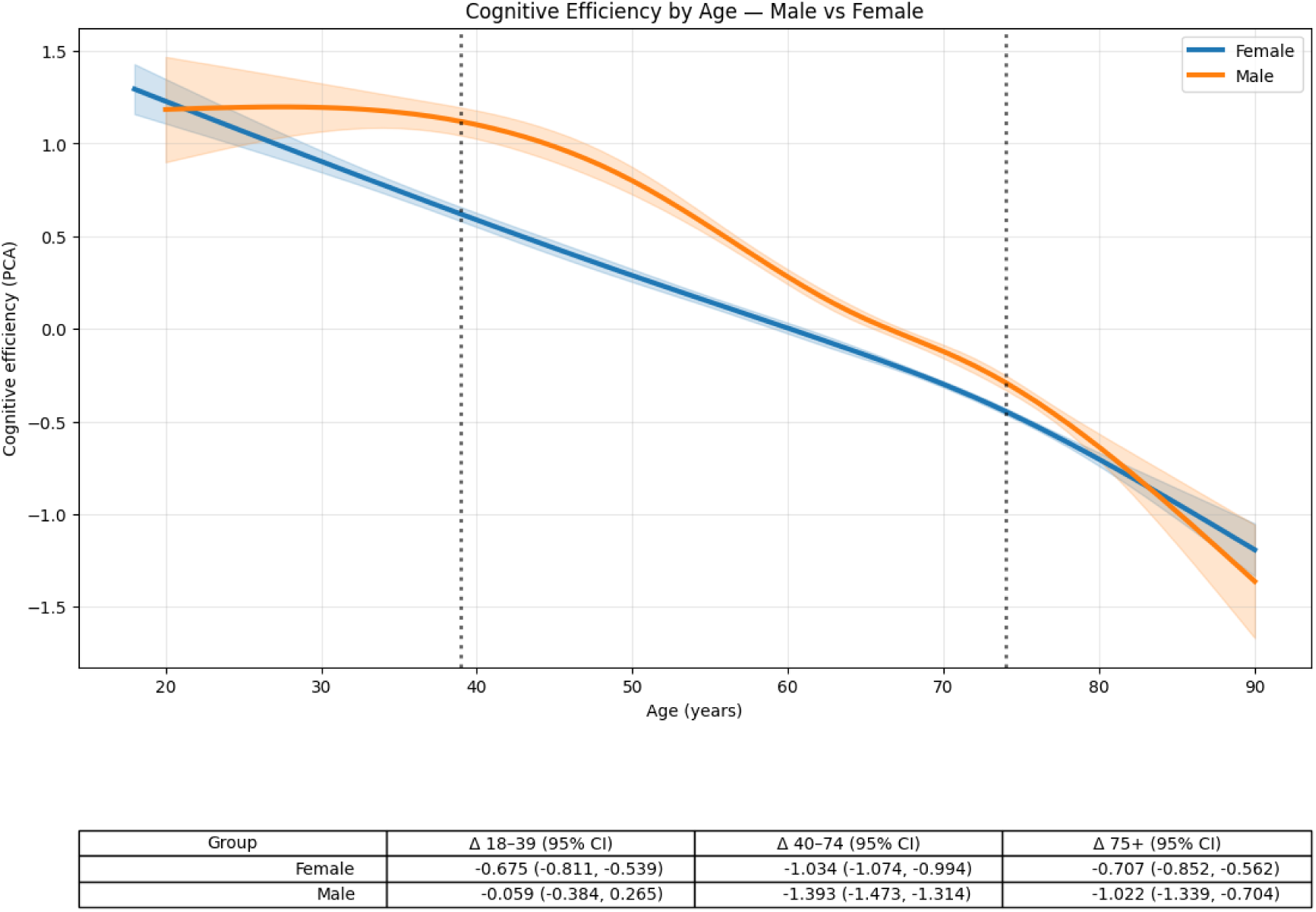
Age-related trajectories of cognitive efficiency by sex. Trajectories of cognitive efficiency (PC1) stratified by sex using natural cubic regression splines with 95% confidence intervals. Both groups show declining cognitive efficiency with age, with males showing higher baseline performance but faster decline. Vertical dashed lines indicate spline breakpoints.

### Smoking

Smoking status showed a progressive dose-response relationship with cognitive efficiency (Figure 4). Compared with never-smokers, participants smoking 11–20 cigarettes/day exhibited a β of −0.112 (p = 0.001), corresponding to approximately 3.3 age-equivalent years of additional cognitive aging, while those smoking > 20/day showed β = −0.167 (p < 0.001), equivalent to approximately 5.0 additional years. Light smoking (≤ 10/day) was not significantly associated with cognitive efficiency (p = 0.151).

**Figure 4.**
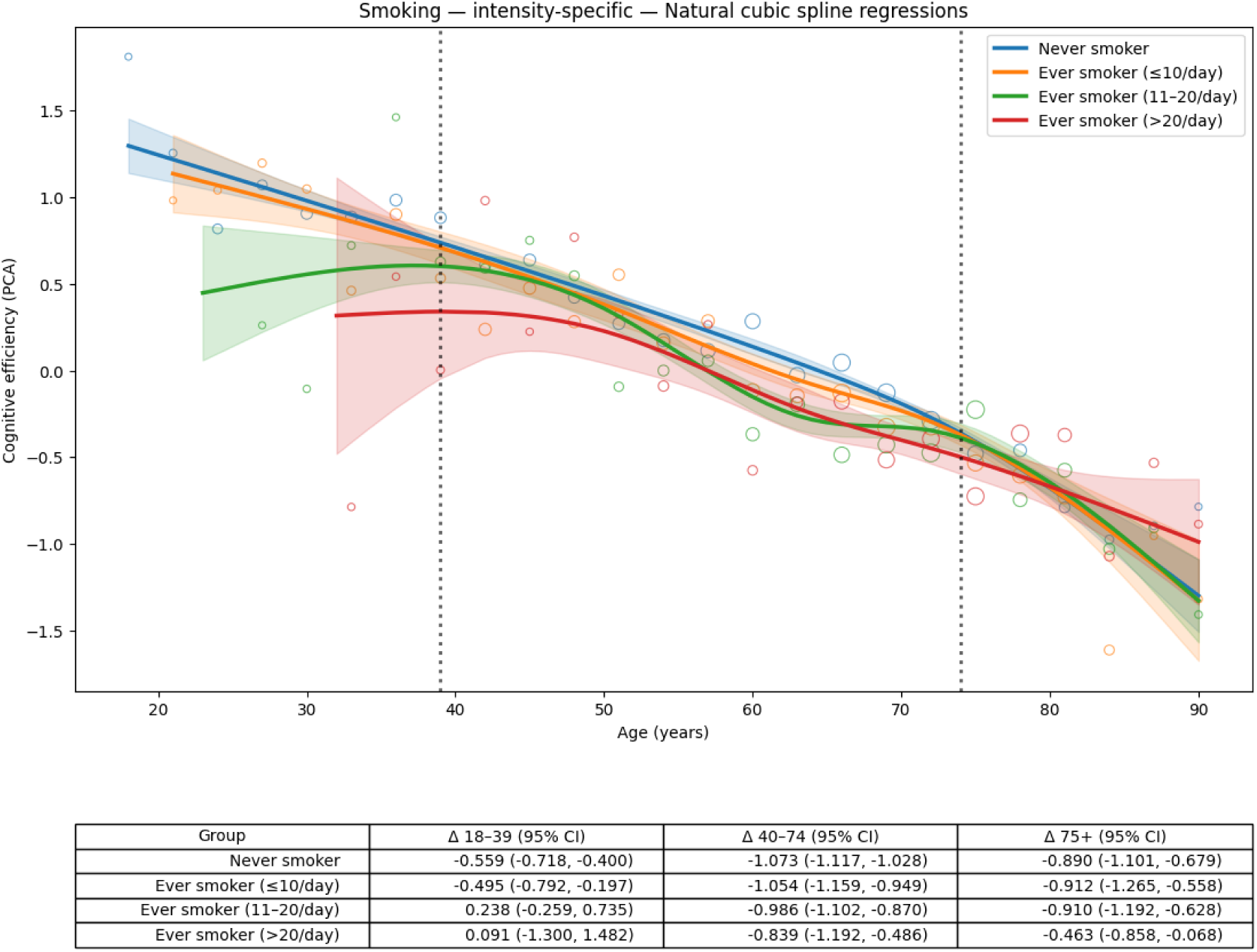
Cognitive efficiency by age and smoking intensity. Cognitive efficiency by age stratified by smoking intensity using natural cubic regression splines with 95% confidence intervals. Heavier smoking is associated with lower cognitive efficiency across adulthood, with differences becoming more pronounced at midlife and older ages.

### Alcohol binge drinking

Binge drinking frequency showed limited associations with cognitive efficiency (Figure 5). Using never-bingeing as the reference, infrequent binge drinking (less than monthly) was significantly associated with a modest decrement (β = −0.063, p = 0.001; +1.9 age-equivalent years).

**Figure 5.**
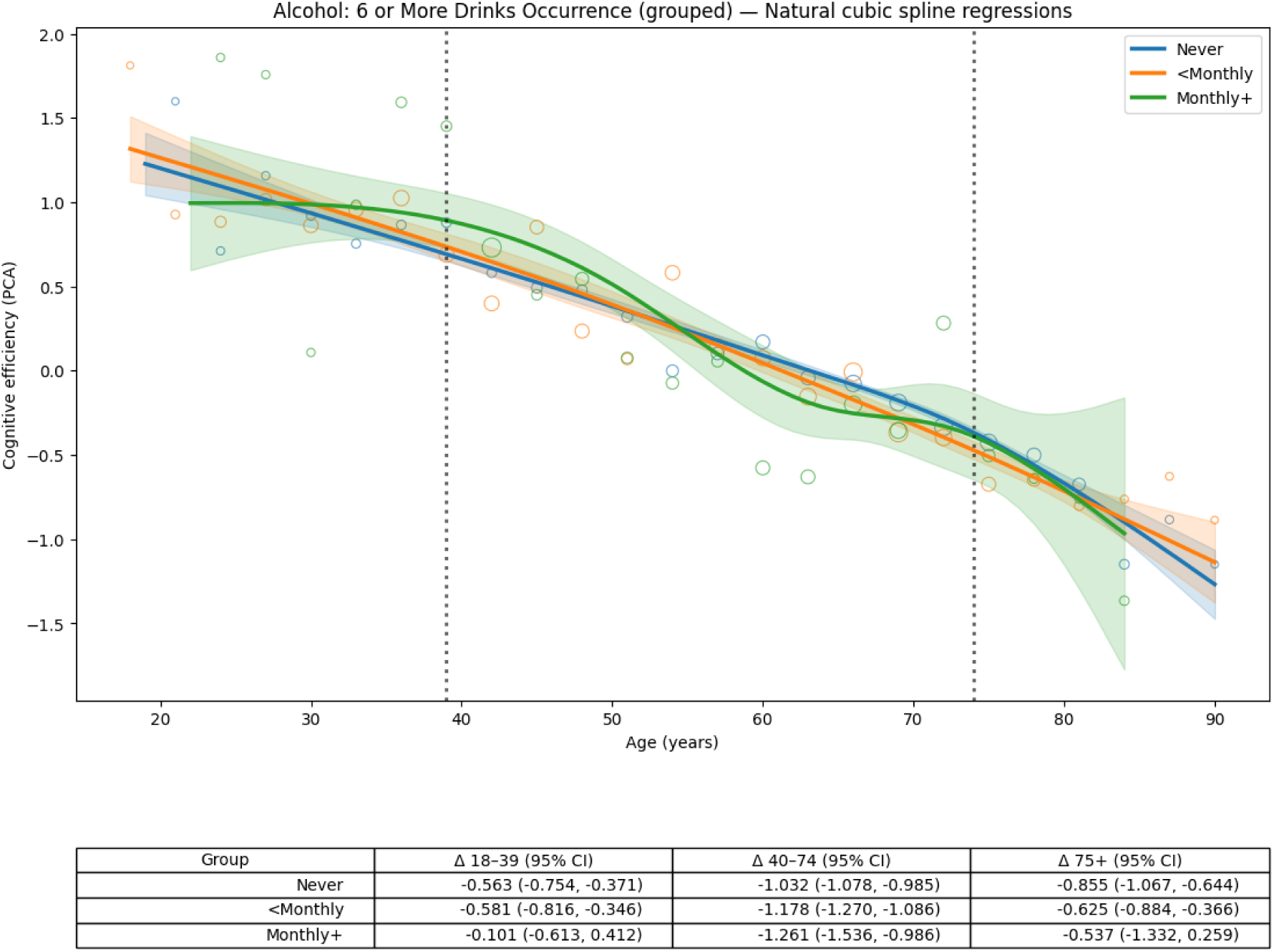
Cognitive efficiency by age and binge drinking frequency. Cognitive efficiency by age stratified by binge drinking frequency (never, less than monthly, monthly or more) using natural cubic spline regression with 95% confidence intervals.

Monthly or more frequent categories were not significantly associated with cognitive efficiency, likely because of small subgroup sizes, survivor bias (heavier drinkers may not reach older ages) and abstainer bias (former heavy drinkers classified as never-bingeing after cessation).

### Social satisfaction

Social satisfaction was the strongest lifestyle predictor of cognitive efficiency (Figure 6). Relative to high satisfaction, moderate satisfaction corresponded to β = −0.124 (p = 0.007; +3.6 age-equivalent years) and low satisfaction to β = −0.192 (p = 0.001; +5.8 age-equivalent years). These findings are consistent with James et al. [9], who reported that highly socially active older adults exhibited nearly 70% slower cognitive decline. Although social activity is a separate construct from social satisfaction, it captures the social dimension within the risk factors available in this dataset. The pattern of increasing cognitive deficit with decreasing social satisfaction was consistent across all age groups.

**Figure 6.**
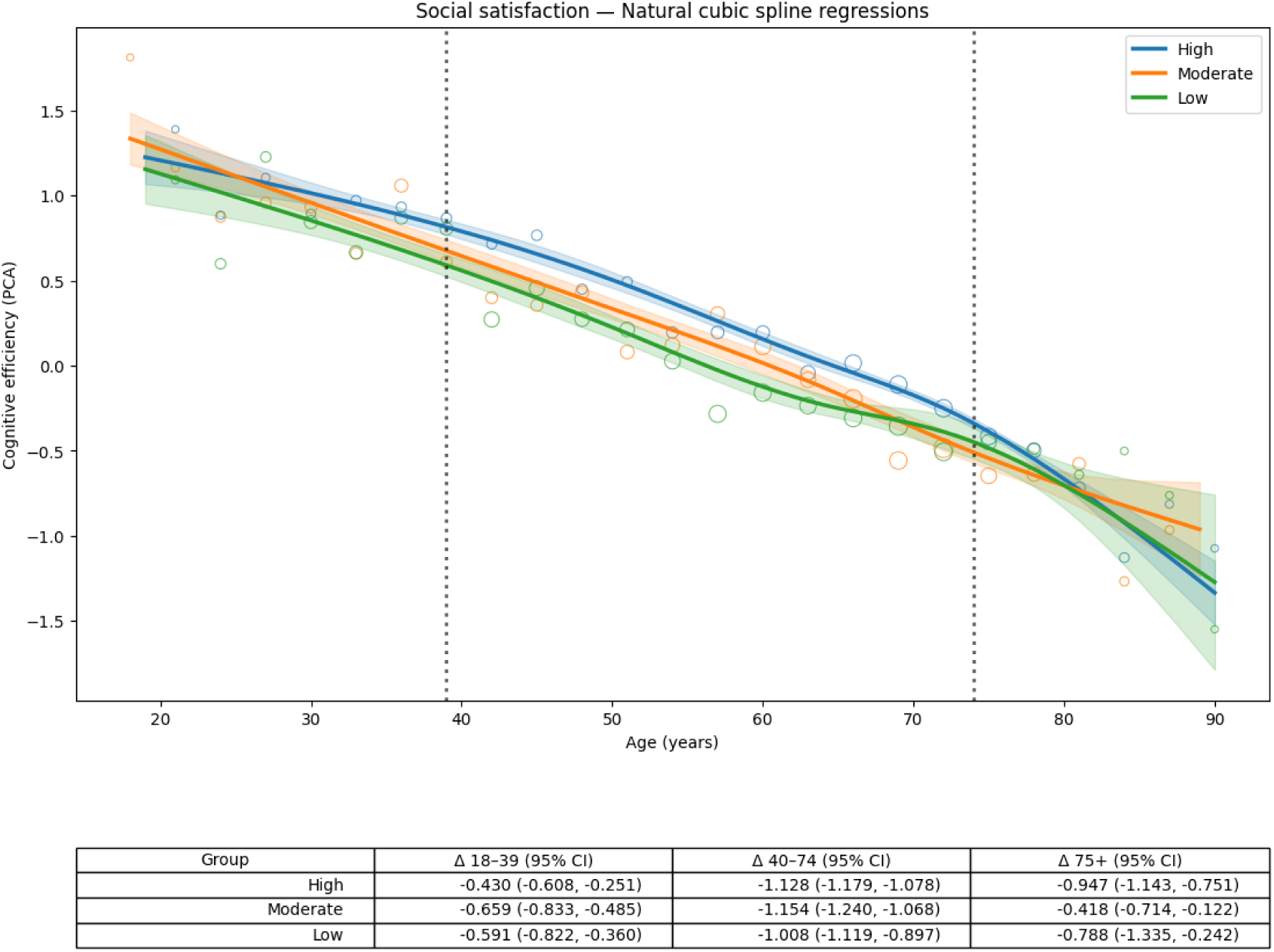
Cognitive efficiency by age and social satisfaction. Cognitive efficiency by age stratified by social satisfaction level (high, moderate, low) using natural cubic spline regression with 95% confidence intervals. Lower social satisfaction is associated with consistently lower cognitive efficiency across adulthood.

### Blood pressure

Blood pressure associations with cognitive efficiency were small and mostly non-significant (Figure 7). Elevated blood pressure (β = 0.019, p = 0.392) and stage 1 hypertension (β = 0.009, p = 0.644) showed small, non-significant positive associations relative to normal blood pressure, consistent with reported U-shaped associations between blood pressure and dementia risk in older adults [15]. Stage 2 hypertension showed a modest negative association (β = −0.038, p = 0.084; +1.1 age-equivalent years) that did not reach statistical significance.

**Figure 7.**
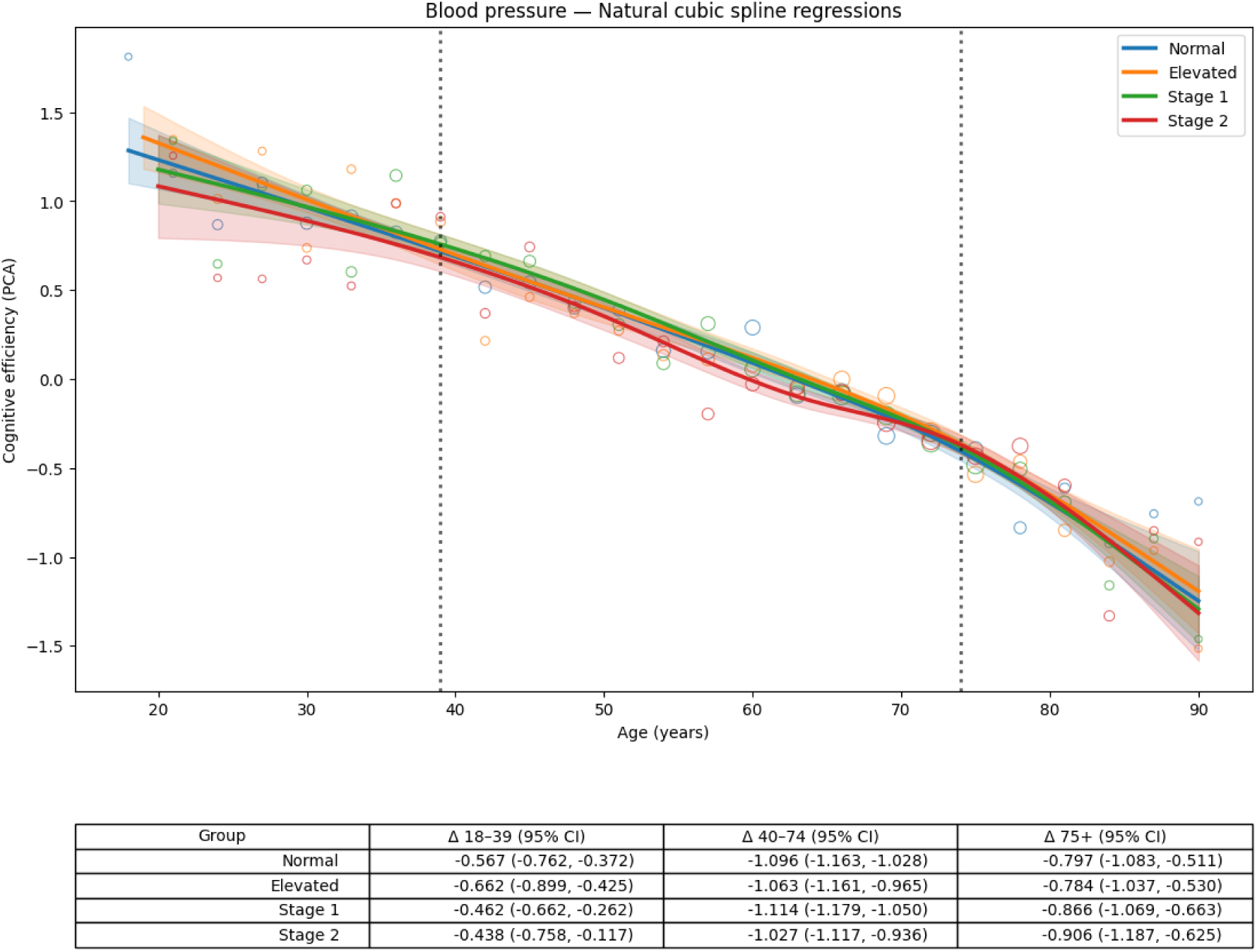
Cognitive efficiency by age and blood pressure category. Cognitive efficiency by age stratified by blood pressure category (normal, elevated, stage 1 and stage 2 hypertension) using natural cubic spline regression with 95% confidence intervals. Modest differences are observed across hypertension stages.

### Path analysis

Single-outcome path analysis confirmed that low social satisfaction (β = −0.194, p < 0.001) and heavy smoking (β = −0.167, p < 0.001) were the strongest lifestyle-related predictors of cognitive efficiency, exceeding the magnitude of alcohol and blood pressure effects (R² = 0.264; Figure 8). The multi-outcome path model further showed that smoking and low social satisfaction were consistently associated with lower d′ and slower median reaction time, indicating broad effects on attentional processing rather than on sensitivity alone.

**Figure 8.**
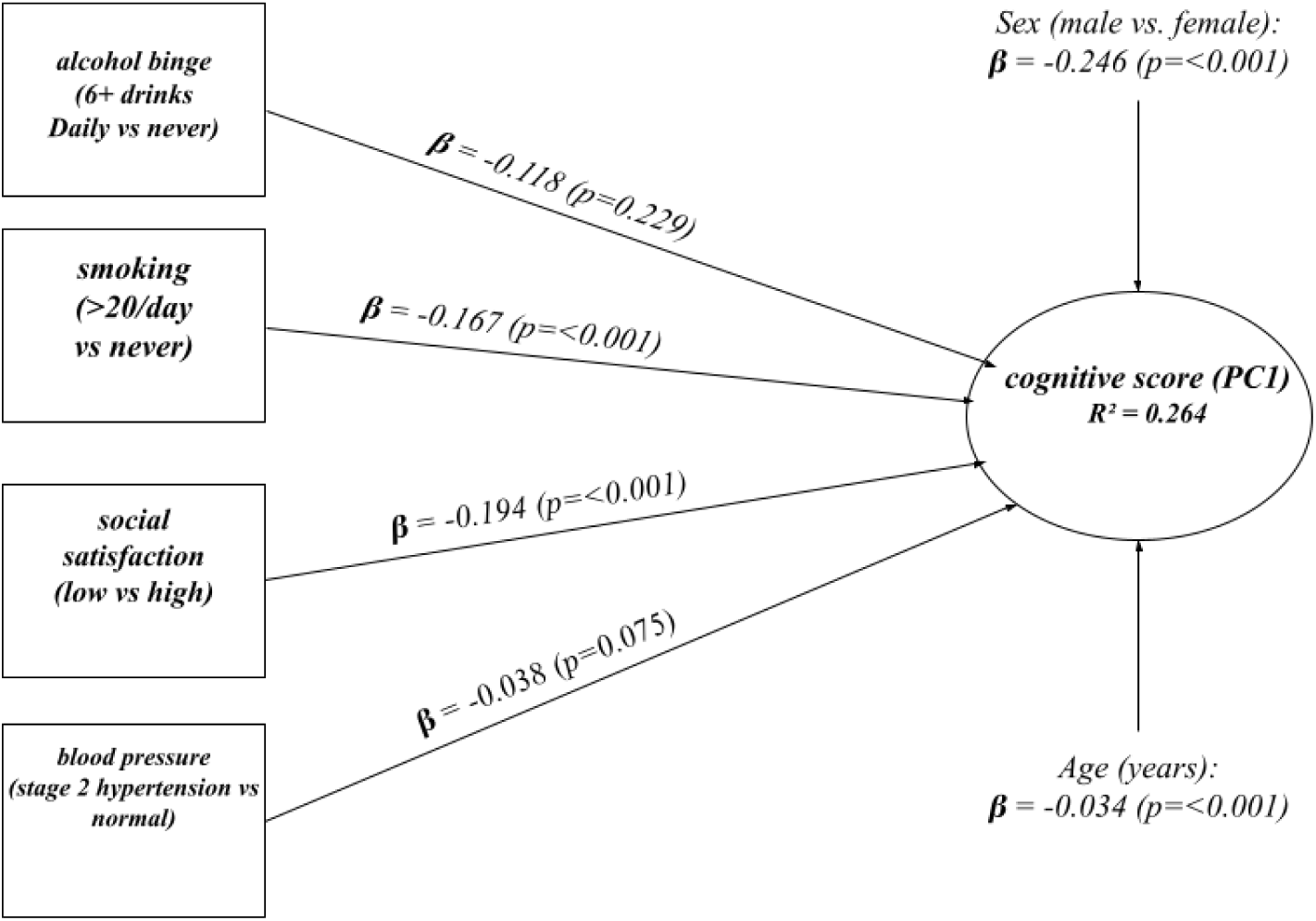
Path diagram of standardised predictors of cognitive efficiency. Standardised regression coefficients (β) predicting cognitive efficiency (PC1) from age, sex, smoking, social satisfaction, alcohol binge drinking and blood pressure. Social satisfaction and smoking show the largest lifestyle-related associations.

### Combined risk factors

Bootstrapped analyses (5000 resamples) showed that combinations of risk factors produced cognitive deficits substantially larger than those observed for single factors (Table 4). Among statistically significant multi-factor contrasts, the most pronounced associations involved low social satisfaction in combination with other risk factors: monthly-or-more binge drinking combined with low social satisfaction (+6.9 years, 95% bootstrap CI +2.2 to +11.8; p = 0.004), heavy smoking combined with low social satisfaction and high blood pressure (+7.5 years, 95% bootstrap CI +2.9 to +11.9; p = 0.002), and monthly-or-more binge drinking combined with low social satisfaction and high blood pressure (+7.8 years, 95% bootstrap CI +2.7 to +12.9; p = 0.004). The four-factor combination (heavy smoking, monthly-or-more binge drinking, low social satisfaction and stage 2 hypertension) corresponded to +12.9 age-equivalent years (95% bootstrap CI +3.5 to +25.0; p = 0.014), although this estimate was based on fewer than 20 observed individuals and warrants cautious interpretation. Individual risk factors that were non-significant or modest in isolation, particularly binge drinking and elevated blood pressure, showed substantially larger associations when co-occurring with low social satisfaction, suggesting that social dissatisfaction may amplify cognitive vulnerability to co-occurring behavioral and vascular risks.

**Table 4.** Selected bootstrapped combined risk profiles at age 50.

| **Risk profile** | **n** | **p (bootstrap)** | **Age-equivalent years (95% CI)** |
| --- | --- | --- | --- |
| No risk factors (reference) | 5504 | Ref | 0 (ref) |
| Low social satisfaction only | 780 | < 0.001 | +2.6 (+1.2 to +4.1) |
| Heavy smoking only | 210 | < 0.001 | +5.2 (+2.8 to +7.5) |
| High blood pressure only | 5357 | 0.346 | +0.3 (−0.4 to +1.1) |
| Low social satisfaction + high blood pressure | 774 | < 0.001 | +4.5 (+3.0 to +6.0) |
| Binge drinking + low social satisfaction | 50 | 0.004 | +6.9 (+2.2 to +11.8) |
| Heavy smoking + low social satisfaction + high blood pressure | 50 | 0.002 | +7.5 (+2.9 to +11.9) |
| Binge drinking + low social satisfaction + high blood pressure | 59 | 0.004 | +7.8 (+2.7 to +12.9) |
| All four risk factors combined | < 20 | 0.014 | +12.9 (+3.5 to +25.0) |

Estimates are model predictions anchored at age 50, derived from 5000 bootstrap resamples. Heavy smoking, > 20 cigarettes/day; binge drinking, monthly or more; high blood pressure, stage 2 hypertension. Cell counts below 20 are suppressed per the All of Us Data and Statistics Dissemination Policy. CI, confidence interval; Ref, reference category.

## Discussion

This study examined associations between modifiable lifestyle risk factors and GradCPT-based sustained visual attention performance across adulthood in a large, nationally diverse sample. Three hypotheses were tested and broadly supported.

First, sustained attention performance varied substantially across the adult lifespan, peaking around age 40 and declining steadily thereafter, consistent with Fortenbaugh et al. [7].

Expressing effect sizes as age-equivalent years places lifestyle-associated differences in a clinically intuitive context. Second, modifiable risk factors showed independent associations with cognitive efficiency after adjustment for age and sex. Social satisfaction was the strongest lifestyle predictor, consistent with evidence that socially engaged individuals maintain higher cognitive reserve [9, 16]. Smoking showed a dose-response pattern aligning with structural evidence of smoking-accelerated brain aging [8]. Blood pressure associations were modest and inconsistent, possibly reflecting U-shaped relationships between blood pressure and cognition in older adults, in whom higher blood pressure may be protective at older ages [15]. Alcohol binge drinking showed limited significant associations, likely complicated by survivor and abstainer biases inherent in cross-sectional designs.

Third, combined risk factor models revealed synergistic effects. Several behaviours that were individually non-significant, notably binge drinking and elevated blood pressure, showed substantially larger deficits when co-occurring with low social satisfaction, suggesting an interactive rather than an additive model of cognitive risk. This finding is consistent with Bittner et al. [12], who demonstrated that cumulative lifestyle risk scores are associated with accelerated structural brain aging beyond the sum of individual contributions, and with prior evidence that social and vascular pathways may share harmful mechanisms, including inflammation, neurotoxicity and psychosocial stress [17].

Sex emerged as a major non-modifiable differentiator, with males showing higher baseline performance but faster decline, converging with female performance by late adulthood, consistent with prior visuospatial cognition literature [14]. While not the primary focus, this reinforces the importance of demographic covariate adjustment in future cognitive aging models.

### Limitations

The cross-sectional design limits causal inference: longitudinal follow-up is required to determine whether observed differences predict future cognitive decline at the individual level. Lifestyle measures were self-reported and subject to recall and social desirability bias. Although the All of Us dataset is more diverse than many prior samples, it over-represents older adults and certain demographic groups (White, female, mean age 61.1 years). Subgroup cell sizes in higher-order combined-exposure analyses were small, warranting cautious interpretation of extreme estimates from the bootstrapped analysis. Cognitive outcomes are limited to GradCPT-based sustained attention and do not capture broader functional impairment or clinical diagnoses.

## Conclusions

Modifiable lifestyle and psychosocial factors are meaningfully associated with age-adjusted performance on the GradCPT, a behavioral measure of sustained visual attention and cognitive control. Beyond identifying individual risk factors, the analysis indicates that specific combinations of behaviours, particularly those involving low social satisfaction, are associated with substantially larger cognitive deficits than would be expected from cumulative effects alone.

Expressing these associations in age-equivalent units provides an interpretable framework for understanding how everyday behaviours may relate to cognitive aging. These results support an integrated, prevention-focused approach to cognitive health and warrant longitudinal studies to establish causal pathways and identify optimal intervention targets.

## Supporting information

IRB Exemption Letter

## Ethics statement

Data were obtained from the All of Us Research Program, which operates under National Institutes of Health Institutional Review Board oversight. All participants provided electronic informed consent prior to enrolment in the All of Us Research Program, including consent for the secondary research use of their de-identified data. The present secondary analysis of de-identified registered-tier data was reviewed by the Morgan State University Institutional Review Board and determined to be exempt from full review.

## Data availability

The datasets analysed during the current study are available through the All of Us Research Hub (https://workbench.researchallofus.org), subject to approval of a data access request and completion of the required training. Individual-level data cannot be redistributed by the authors under the terms of the All of Us Data Use and Registration Agreement. Analysis code is available from the corresponding author on reasonable request.

## Author contributions

NCC contributed to conceptualisation, data curation, formal analysis, methodology and writing of the original draft. TB contributed to guidance on data analysis within the All of Us Dataset, and ARS contributed to methodology, direction, and applying scaling to the analysis. IKT contributed to supervision and to review and edit the manuscript. All authors read and approved the final manuscript.

## Acknowledgements

We gratefully acknowledge All of Us participants for their contributions, without whom this research would not have been possible. We also thank the National Institutes of Health’s All of Us Research Program for making available the participant data examined in this study. The All of Us Research Program is supported by the National Institutes of Health.

We would also like to acknowledge Dr. Fahmi Khalifa Assistant Professor, Department of Electrical and Computer Engineering at Morgan State University and Dr. Walter Royal III Director, Center for Brain Science, Biology at Morgan State University for providing feedback throughout the manuscript process.

This study was reviewed and approved by Morgan State University’s Ethics Committee under IRB #26/10-0268.

## Use of large language models and AI tools

During manuscript preparation, the authors used Claude (Anthropic) for language editing and copy-editing assistance. The tool was not used to generate original content, data, analyses or interpretations. The authors reviewed the output and take full responsibility for the content of this article.

## Abbreviations

CI: confidence interval
CV: coefficient of variation
DBP: diastolic blood pressure
GradCPT: Gradual-Onset Continuous Performance Task
NIH: National Institutes of Health
OLS: ordinary least squares
PC1: first principal component
PCA: principal component analysis
PROMIS: Patient-Reported Outcomes Measurement Information System
RT: reaction time
SBP: systolic blood pressure
SD: standard deviation
SE: standard error

