## Supplementary material for "Social and behavioral risk factors are associated with age-related differences in sustained attention: A cross-sectional GradCPT analysis of the All of Us dataset": IRB Exemption Letter

MORGAN STATE UNIVERSITY

GROWING THE FUTURE ● LEADING THE WORLD

**Institutional Review Board (IRB)**

October 22, 2026

Morgan State University
IRB
**EXEMPT**
**Signature:** Benjamin Welsh
**Date:** 10/23/2026
**No further action needed.**

Dr. Ingrid K. Tulloch
College of Liberal Arts
Morgan State University

RE: IRB #26/10-0268

Dear Dr. Ingrid K. Tulloch,

Following administrative review of the materials submitted to the Institutional Review Board (IRB) for the study in collaboration with Natan Coresh-Chiappori titled *"The Relationship Between Lifestyle Risk Factors and Cognitive Efficiency Across Age: A cross-sectional analysis of the All of Us dataset",* the project has been determined to be exempt from IRB review under 45 CFR §46.104(d) Category 4: Secondary Research Use of Identifiable Private Information or Biospecimens of the Common Rule. This exemption applies to research involving Secondary Research Use of Identifiable Private Information or Biospecimens**.**

Please note the following Principal Investigator responsibilities:

- Notify the IRB prior to implementing any substantive modifications to the study, as changes may affect the exemption determination under 45 CFR §46.104.
- Notify the IRB when all research activities have concluded so the protocol may be administratively closed.

If you have any questions, please contact or the IRB at.

Sincerely,

Benjamin Welsh, Ph.D.
IRB Chairperson
Cc: Dr. Keyshawn Moncrieffe IRB Administrator
