## Supplemental Files for "Social and behavioral risk factors are associated with age-related differences in sustained attention: A cross-sectional GradCPT analysis of the All of Us dataset"

*Social and behavioral risks accelerate attentional aging: a cross-sectional GradCPT analysis of the All of Us dataset*

**Supplementary Table S1.** Full multiple linear regression of cognitive efficiency (PC1), including race/ethnicity coefficients

| Risk factor | β (SE) | t | p value |
|---|---|---|---|
| Age (years) | −0.035 (0.001) | −63.234 | < 0.001 |
| Sex: male | Ref | — | — |
| Sex: female | −0.241 (0.016) | −14.985 | < 0.001 |
| Race: non-Hispanic White | Ref | — | — |
| Race: non-Hispanic Black | −0.411 (0.039) | −10.658 | < 0.001 |
| Race: Hispanic | −0.241 (0.030) | −8.050 | < 0.001 |
| Race: Asian | −0.006 (0.048) | −0.134 | 0.893 |
| Race: other/multiracial | −0.116 (0.033) | −3.505 | < 0.001 |
| Race: unknown | −0.000 (0.000) | −1.329 | 0.184 |
| Smoking: never smoker | Ref | — | — |
| Smoking: ever smoker (≤ 10/day) | −0.028 (0.020) | −1.375 | 0.169 |
| Smoking: ever smoker (11–20/day) | −0.124 (0.023) | −5.391 | < 0.001 |
| Smoking: ever smoker (> 20/day) | −0.183 (0.035) | −5.280 | < 0.001 |
| Binge drinking: never | Ref | — | — |
| Binge drinking: less than monthly | −0.062 (0.019) | −3.269 | 0.001 |
| Binge drinking: monthly | 0.003 (0.037) | 0.079 | 0.937 |
| Binge drinking: weekly | −0.015 (0.057) | −0.269 | 0.788 |
| Binge drinking: daily | −0.093 (0.097) | −0.955 | 0.340 |
| Social satisfaction: high | Ref | — | — |
| Social satisfaction: moderate | −0.114 (0.017) | −6.585 | < 0.001 |
| Social satisfaction: low | −0.172 (0.023) | −7.510 | < 0.001 |
| Blood pressure: normal | Ref | — | — |
| Blood pressure: elevated | 0.019 (0.022) | 0.858 | 0.391 |
| Blood pressure: stage 1 hypertension | 0.010 (0.018) | 0.541 | 0.589 |
| Blood pressure: stage 2 hypertension | −0.026 (0.021) | −1.256 | 0.209 |

N = 13,866; $R^2$ = 0.273. PC1, first principal component; Ref, reference category; SE, standard error.
